# Deep Learning Model for Improving the Characterization of Coronavirus on Chest X-ray Images Using CNN

**DOI:** 10.1101/2020.10.30.20222786

**Authors:** Olaide N. Oyelade, Absalom E. Ezugwu

**Affiliations:** School of Mathematics, Statistics, and Computer Science, University of KwaZulu-Natal, King Edward Avenue, Pietermaritzburg Campus, Pietermaritzburg, 3201, KwaZulu-Natal, South Africa; Department of Computer Science, Ahmadu Bello University Zaria

**Keywords:** Coronavirus, COVID-19, machine learning, deep learning, convolutional neural network, CNN, X-ray

## Abstract

The novel Coronavirus, also known as Covid19, is a pandemic that has weighed heavily on the socio-economic affairs of the world. Although researches into the production of relevant vaccine are being advanced, there is, however, a need for a computational solution to mediate the process of aiding quick detection of the disease. Different computational solutions comprised of natural language processing, knowledge engineering and deep learning have been adopted for this task. However, deep learning solutions have shown interesting performance compared to other methods. This paper therefore aims to advance the application deep learning technique to the problem of characterization and detection of novel coronavirus. The approach adopted in this study proposes a convolutional neural network (CNN) model which is further enhanced using the technique of data augmentation. The motive for the enhancement of the CNN model through the latter technique is to investigate the possibility of further improving the performances of deep learning models in detection of coronavirus. The proposed model is then applied to the COVID-19 X-ray dataset in this study which is the National Institutes of Health (NIH) Chest X-Ray dataset obtained from Kaggle for the purpose of promoting early detection and screening of coronavirus disease. Results obtained showed that our approach achieved a performance of 100% accuracy, recall/precision of 0.85, F-measure of 0.9, and specificity of 1.0. The proposed CNN model and data augmentation solution may be adopted in pre-screening suspected cases of Covid19 to provide support to the use of the well-known RT-PCR testing.

## 1 Introduction

The 2019 novel coronavirus disease presents an important and urgent threat to global health, and has equally exposed to an extent the fragility of the most highly placed health institutions and infrastructures across the globe [27, 28]. Since the first status recorded case of COVID-19 was identified in early December 2019 in Wuhan, in the Hubei province of the People’s Republic of China, the number of patients confirmed to have contracted the disease has exceeded 35,960,908 in 214 countries, and the number of people infected is probably much higher [29]. Moreover, a record estimate of more than 1,052,310 people have died from the coronavirus COVID-19 outbreak as of October 06, 2020 [29]. However, despite several public health responses aimed at containing the disease and delaying its spread, many countries have now been confronted with critical health care catastrophes ranging from limited hospital beds, shortage of medical equipment, and contamination of medical frontline workers [27].

In order to alleviate the liability placed on the already fragile healthcare system, while also providing the best possible medical care for patients, efficient diagnosis and treatments of the novel coronavirus disease is urgently needed. The study conducted by Wynants et al. [27] revealed that the development of efficient prediction models that combine several variables or features to estimate the risk of people being infected by the disease or experiencing a poor outcome from the infection could assist medical staff in triaging patients when allocating limited healthcare resources [27]. Several advanced artificial intelligence and machine learning models, specifically deep learning algorithms, have been proposed as evidenced in many academic databases and journals in response to a call by the WHO to share relevant COVID-19 research findings rapidly and openly to inform the public health response and help save people’s lives.

Singh et al. [32, 34] developed a deep convolution neural network (CNN) that was applied in the automated diagnosis and analysis of COVID-19 in infected patients. The authors’ proposed model involved the tuning of hyper-parameters of the CNN model with a multi-objective adaptive differential evolution algorithm. Results from the comparative analysis showed that their proposed method outperformed existing machine learning models such as CNN, GA-based CNN, and PSO based CNN, based on the different performance metrics that were employed to validate the conducted experiment, such as the F-measure, Sensitivity, Specificity, and Kappa statistics. Jaiswal et al. [33] developed DenseNet201, a deep transfer machine learning model for the diagnosis and detection of COVID-19 cases from chest (Computed Tomography) CT scans. The proposed model was also utilized to extract some features by adopting its own learned weights on the ImageNet dataset along with a convolutional neural structure. The DenseNet201 model achieved a 97% accuracy compared to other models. In the study presented by Barstugan et al. [35], a machine learning approach was proposed for the early detection of the COVID-19 on abdominal Computed Tomography (CT) images. The authors’ results showed that their model was able to differentiate COVID-19 specific characteristics from other viral pneumonia. Asif and Yi [36] and Asif et al. [37] implemented a deep convolutional neural networks model that was able to automatically detect COVID-19 pneumonia patients using digital chest X-ray radiographs. Hu et al. [38] employed a supervised deep learning model aimed at detecting and classifying COVID-19 infection from CT images and at the same time minimizing the requirements for manual labelling of CT images. Similarly, the model could efficiently distinguish between -ve and +ve cases of COVID-19 by using samples from retrospectively extracted CT images from multi-scanners and multicenter. The experimental results showed that the existing supervised learning model is able to achieve high precision classifications, as well as good qualitative visualization for the lesion detections. Al-antari et al. [39] presented a simultaneous deep learning computer-aided diagnostic tool developed and based on the YOLO predictor for the detection and diagnoses of COVID-19 lung disease from the entire chest X-ray images. Their model was evaluated through five-fold tests for multi-class prediction problem by using two different chest X-ray images. From the experimental results, the infected regions of COVID-19 from the whole X-ray images were simultaneously detected and classified end-to-end through the CAD predictor, which achieved overall detection and classification accuracies greater than 90%. Moreover, the CAD deep learning approach showed greater reliability in assisting health care systems, patients, and physicians to deliver their practical validations. For a comprehensive review of existing machine learning models for COVID-19, interested readers are referred to the following references [30, 31].

Although different artificial intelligence approaches also exist, like the case-based reasoning (CBR) [25] which have been applied to the detection of this disease, CNN methods however have shown to be more effective and promising. Several studies [4, 5, 6, 78, 26, 30] and reviews which have adapted CNN to the task of detection and classification of COVID-19 have proven that the deep learning model is one of the most popular and effective approaches in the diagnosis of COVD-19 from digitized images. This outstanding performance of CNN is due to its ability to learn features automatically from digital images as has been applied to diagnoses of COVID-19 based on clinical images, CT scans, and X-rays of the chest by researchers. Therefore, considering the advantages of the several automated deep learning solution approaches as mentioned above for curbing the spread of COVID-19 through early detection, classification, isolation and treatment of affected persons, it would be worthwhile to further investigate the possibility of developing better and more efficient variants of deep machine learning techniques. Moreover, as revealed in most of the literature implementation results, adapting computational solutions to effectively extract the relevant information inherent in X-Ray imaging can help to automate the process of speeding up diagnoses of SARS-CoV-2 virus.

In this paper, we propose the application of deep learning model in the category of Convolutional Neural Network (CNN) techniques to automate the process of extracting important features and then classification or detection of COVID-19 from digital images, and this may eventually be supportive in overcoming the issue of a shortage of trained physicians in remote communities [24]. The proposed model implementation is such that we first applied some selected image pre-processing techniques to reduce the noise on CT and chest X-rays digital images obtained from the COVID-19 X-ray dataset. All the dataset used to validate the performance superiority of the new model is taken from the National Institutes of Health (NIH) Chest X-Ray datasets. Specifically, the contributions of this research are summarized as follows:

i. Design and development of a new CNN based deep learning framework for the automatic characterization and accurate diagnosis of COVID-19 cases.
ii. The proposed CNN model is aimed at detecting COVID-19 cases using chest X-rays images from a combined COVID-19 X-ray images extracted from the National Institutes of Health (NIH) Chest X-Ray dataset.
iii. Experimentation was done using two optimization algorithms namely Adam and Stochastic gradient descent (SGD).
iv. The implemented CNN based deep learning model was evaluated and its results compared with existing similar state-of-the-art results from literature using the following metrics: accuracy, sensitivity, specificity, F1-score, a confusion matrix, and AUC using receiver operating characteristic (ROC).

The rest of the paper is structured accordingly. In section 2, we explain the proposed deep leaning framework for the characterization of coronavirus on chest X-ray images and datasets. The computational results and different experimentations are reported in section 3. The interpretation of the obtained experimental results from the proposed deep learning model is presented in section 4. Finally, the concluding remarks and future research direction are given in section 5.

## 2 Proposed Approach

In this section, an overview of the deep learning approach proposed in this study is presented. This overview is summarized through an architectural pipeline flow of the concepts comprising it. The datasets and the associated data/image preprocessing techniques adopted for this study are also detailed.

### 2.1 Datasets

The choice and category of datasets for application to any CNN model are very important and require the selection of an appropriate dataset. In this study, we decided to apply our CNN model to chest X-rays or CT images which were outcomes of radiological imaging which have been proven to yield better diagnosis of COVID [1]. Two categories of datasets are employed for the characterization of the features and classification the novel Covid-19 disease. These databases are the COVID-19 X-ray images [2] and the National Institutes of Health (NIH) Chest X-Ray Dataset [3]. The most frequently accessed imaging is the chest X-ray which is due to cost-effectiveness although it presents a more challenging clinical diagnosis task compared with chest CT imaging. Hence, our combined approach of chest X-rays/CT images and the use of publicly available datasets with large instances position our CNN model to achieve clinically relevant diagnoses.

The COVID-19 X-ray dataset consists of cases of Covid-19, MERS, SARS, and ARDS which are all represented as chest X-ray or CT images database. The database is accompanied with several fields for each instance which provides further details on the image sample. These fields include number of days since the start of symptoms or hospitalization of patient (necessary for tracking multiple copies of image taken per patient); sex, age, findings or outcome of the diagnoses, patient survival status, the view of the image presented (PA, AP, or L for X-rays and Axial or Coronal for CT scans), modality (CT or X-ray), clinical notes, and other important information. We obtained 363 instances of images and their accompanied metadata from the COVID-19 X-ray database.

The second database combined with the COVID-19 X-ray dataset in this study is the National Institutes of Health (NIH) Chest X-Ray Dataset. This database is comprised of 112,120 X-ray images which are of sizes 1024 x1024 with disease labels from 30,805 unique patients. The database provides samples of images with their diseases and the disease region bounding boxes. Similar to the COVID-19 X-ray dataset, this database also provides the following metadata about each instance: findings/diagnosis, type of disease diagnosed, age and gender of patient, the view of the image and other details.

In the following figures, we have summarized the class distributions and sizes of the databases and also present a combined chart of the two databases. In Figure 1, we show the number of images in the COVID-19 chest X-ray and NIH Chest X-rays databases which are 363 and 84823 respectively. Figure 2 reveals that the COVID-19 Chest X-ray consists of ten (10) classes of images which include: COVID-19, 287 samples; Streptococcus, 17 samples; ARDS, 16 samples; SARS, 16 samples, Pneumocystis, 15 samples; E.coli, 4 samples, No findings or disease-free images, 3 samples; Chlamydophila, 2 samples; Legionella, 2 samples; and lastly Klebsiella, 1 sample. Similarly, there are 15 classes of images in the NIH Chest X-rays databases (including the ‘No findings or disease-free’ label) which consists of Atelectasis, Consolidation, Infiltration, Pneumothorax, Edema, Emphysema, Fibrosis, Effusion, Pneumonia, Pleural thickening, Cardiomegaly, Nodule Mass and Hernia. The distribution of number of instances across these classes of disease is as follows: No-Finding, 37645 samples; Infiltration, 10814 samples; Effusion, 7567 samples; Atelectasis, 7074 samples; Nodule, 3879 samples; Mass, 3415 samples; Pneumothorax, 2852 samples; Consolidation, 2766 samples; Pleural Thickening, 1984 samples; Cardiomegaly, 1715 samples; Emphysema, 1557 samples; Edema, 1352 samples; Fibrosis, 1219 samples; Pneumonia, 822 samples; and Hernia, 162 samples; These figures are charted in Figure 3.

**Figure 1:**
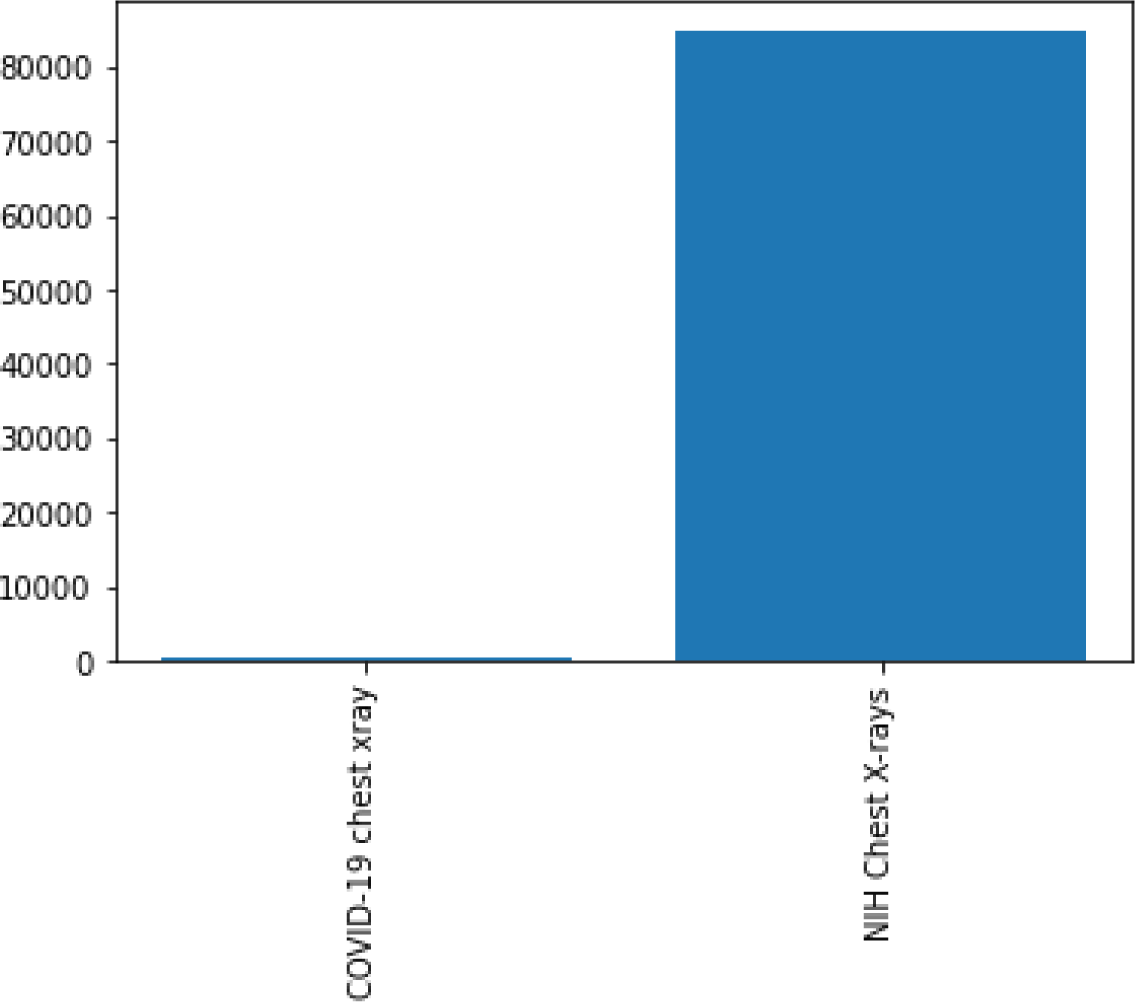
Comparison of the dataset sizes of the two major datasets (Covid-19 and NIH Chest X-rays) used in this study

**Figure 2:**
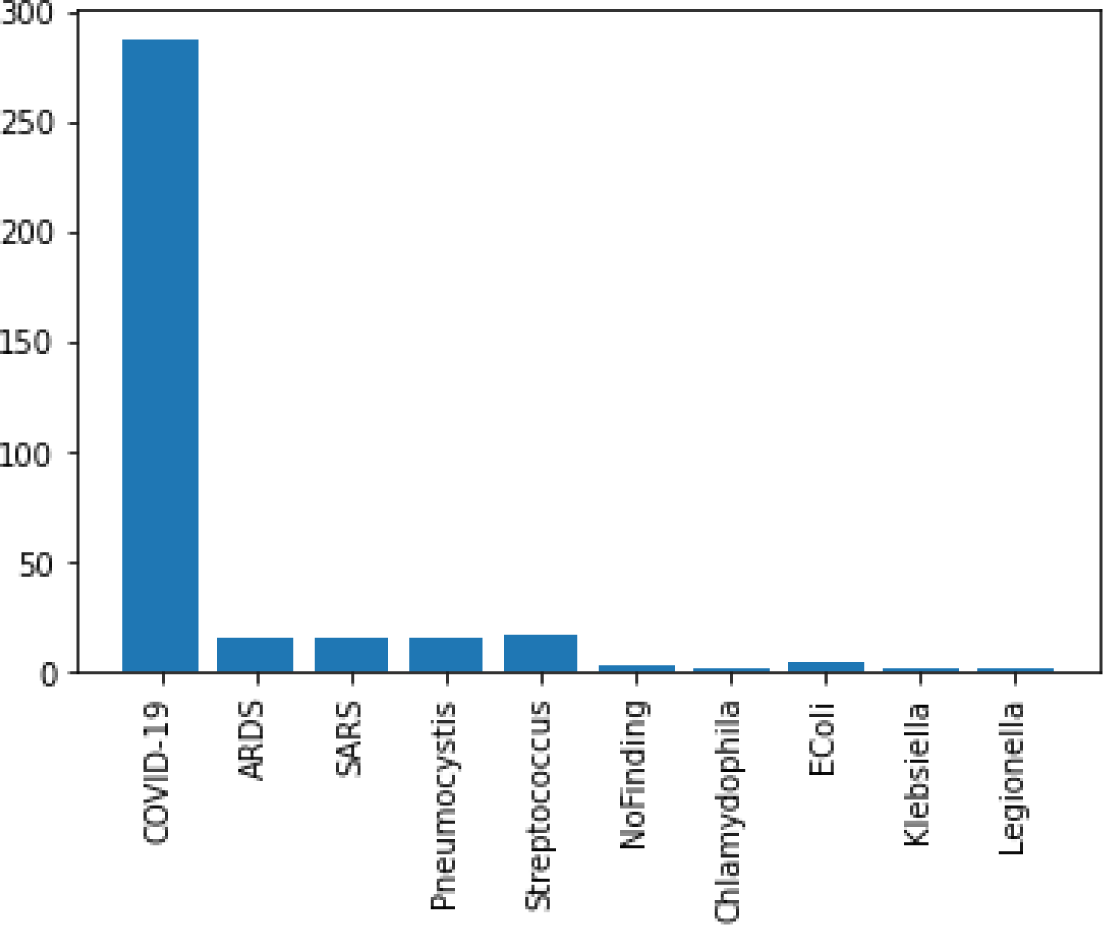
Classes of images available in the Covid-19 Chest X-ray dataset

**Figure 3:**
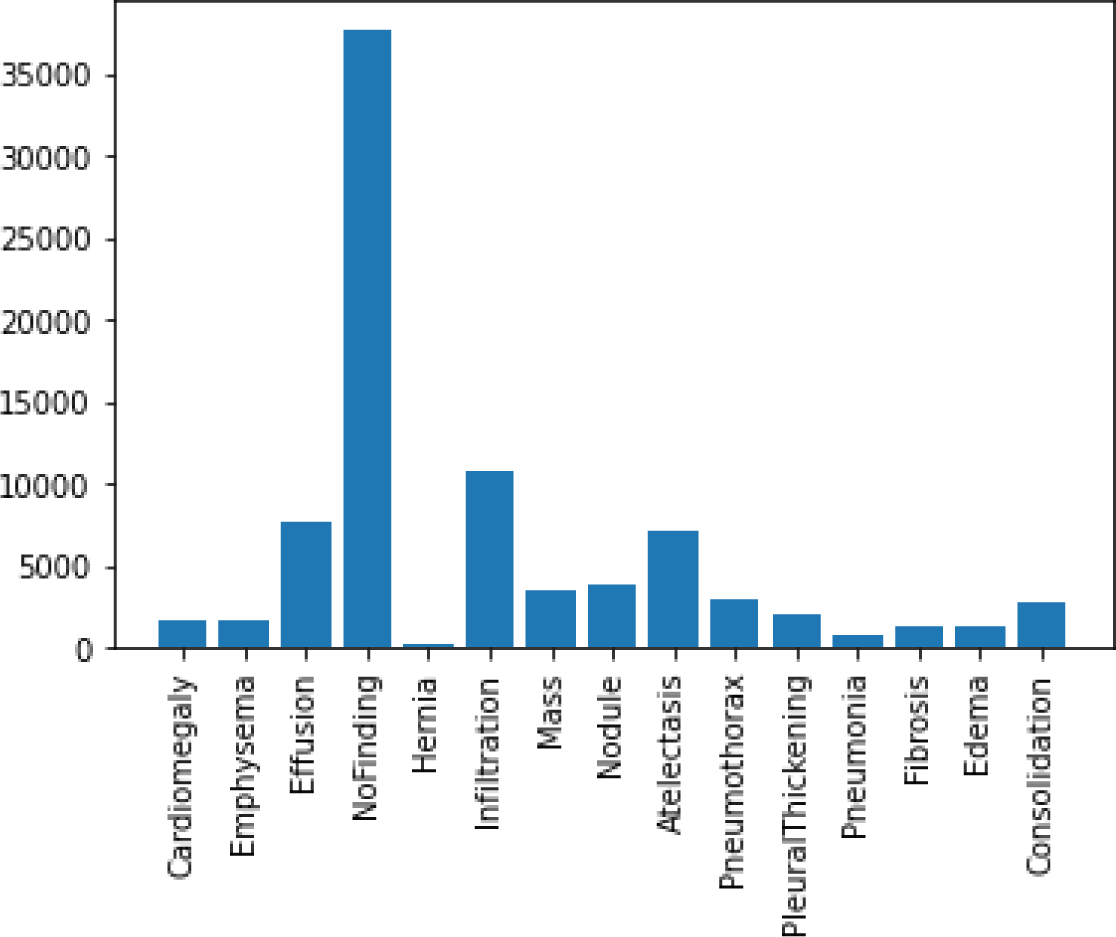
Classes of images available in the NIH Chest X-ray dataset

In the experimentation phase, the combined representation of the images from the two datasets were split into training, evaluation and testing categories and yielded 63887 samples for training, 17034 samples for validation and 4265 samples for testing. This is illustrated in Figure 4. A joint representation of class distribution of images/samples across the two databases for the purpose of training is shown in Figure 5. The combination yielded twenty-four (24) classes with the following number of samples in each class: No-finding or disease free samples has 28222 images, Infiltration has 8017 images, Effusion has 5701 images, Atelectasis has 5373 images, Nodule has 2887 images, Mass has 2558 images, Pneumothorax has 2255 images, Consolidation has 2012 images, Pleural Thickening has 1511 images, Cardiomegaly has 1187 images, Emphysema has 1171 images, Edema has 1003 images, Fibrosis has 968 images, Pneumonia has 636 images, COVID-19 has 203 images, Hernia has 118 images, Streptococcus has 17 images, ARDS has 15 images, Pneumocystis has 15 images, SARS has 11 images, E.coli has 4 images, Chlamydophila has 2 images, Legionella has 2 images, and Klebsiella has 1 image. Meanwhile, a presentation of the splitting of the datasets into evaluation and testing sets are captured in Figures 6 and 7.

**Figure 4:**
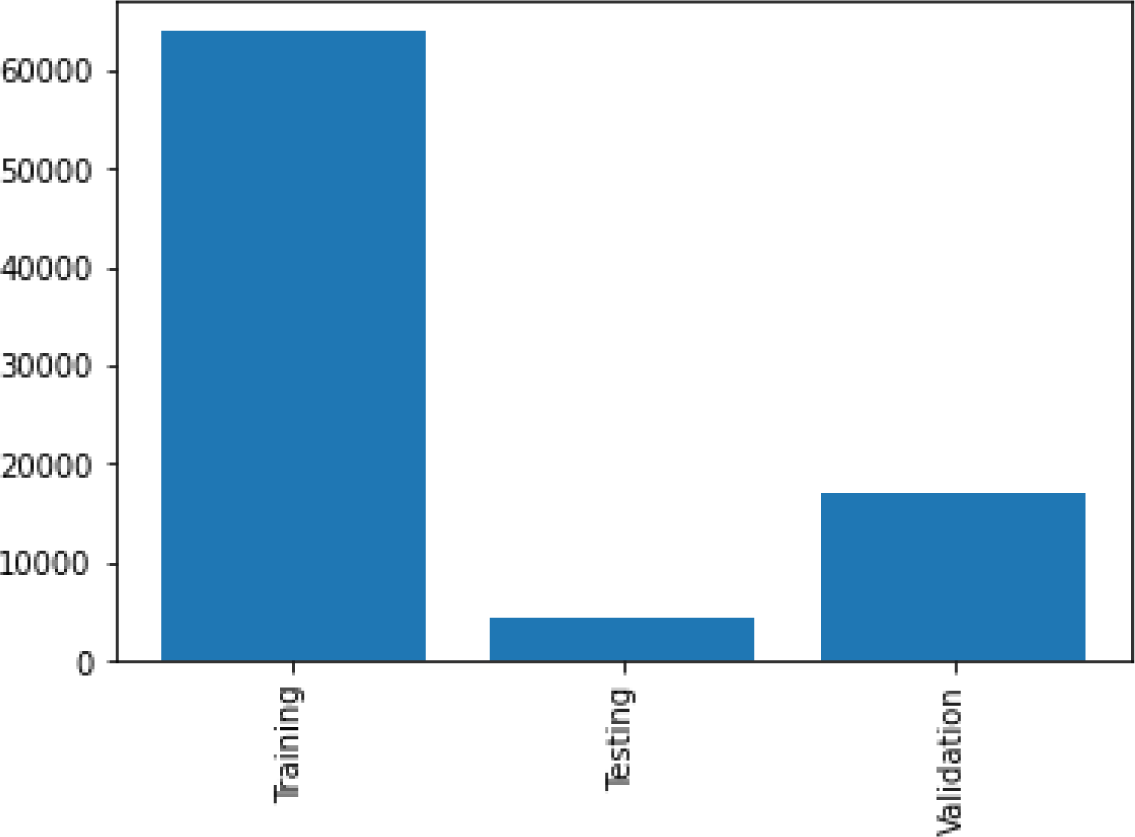
A combine graphing of distribution of images used for training, testing and validation as drawn from the Covid-19 and NIH Chest X-ray datasets

**Figure 5:**
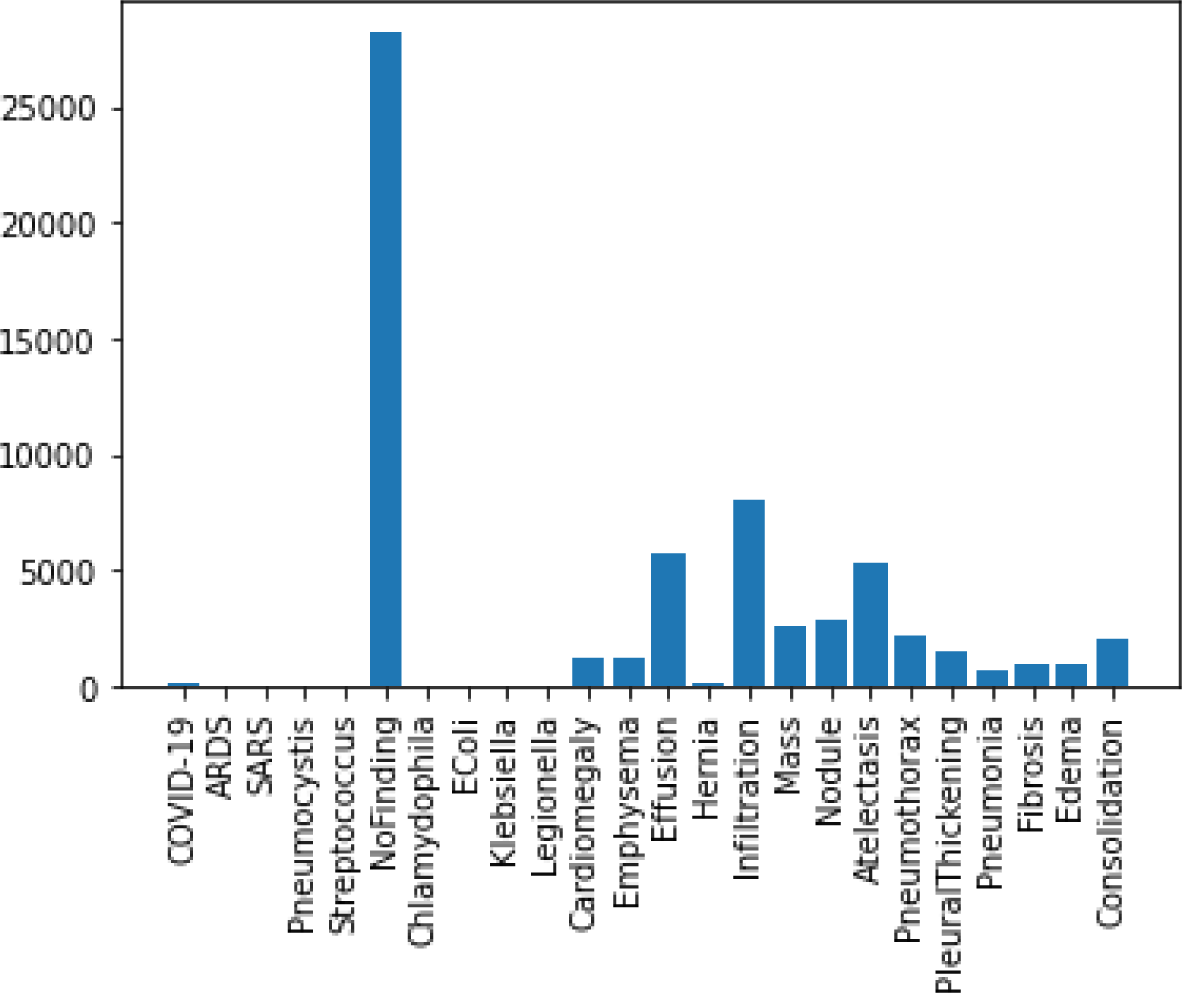
Distribution of training samples among classes of disease as drawn from the Covid-19 Chest X-ray and NIH Chest X-ray datasets

**Figure 6:**
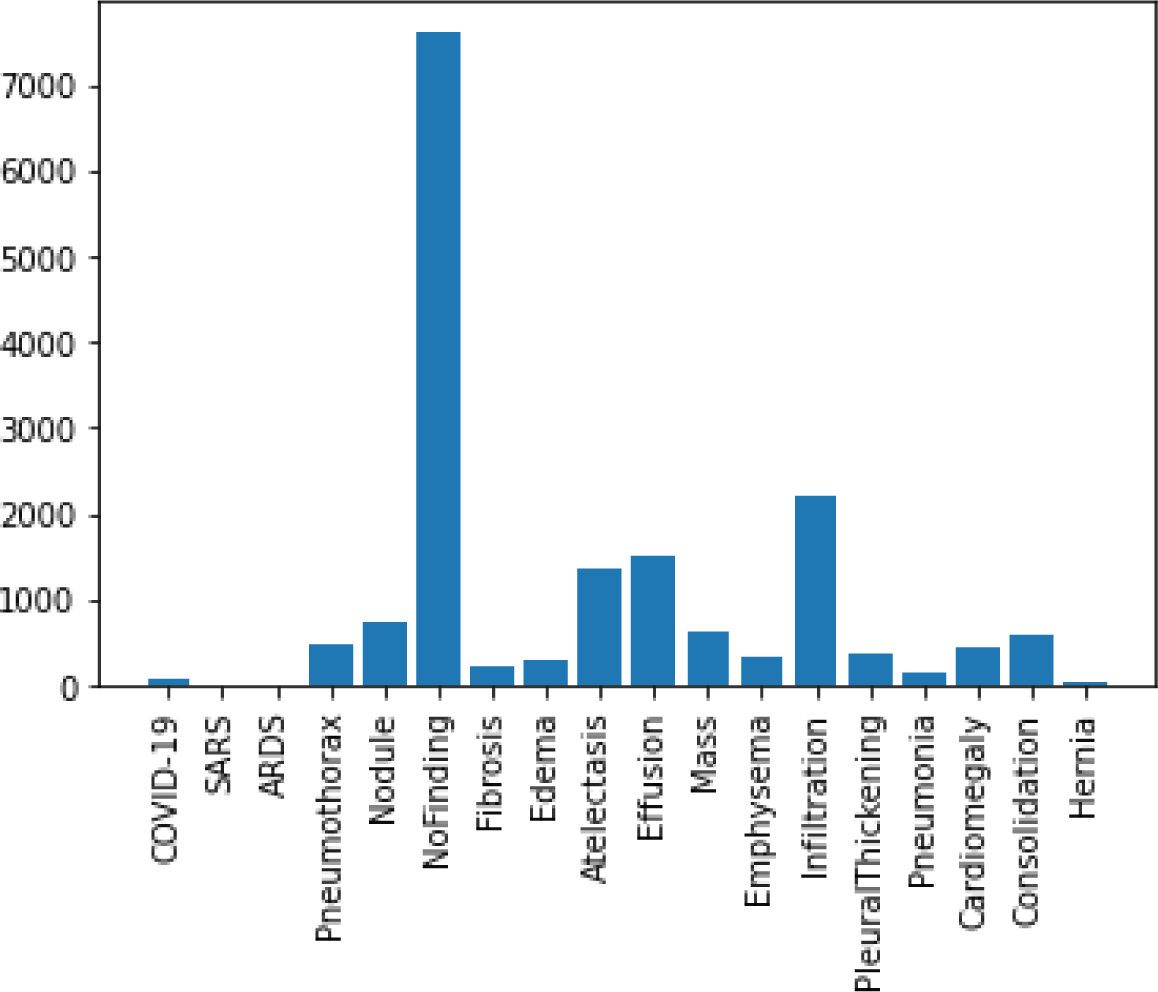
Distribution of validation samples among classes of disease as drawn from the Covid-19 Chest X-ray and NIH Chest X-ray datasets

**Figure 7:**
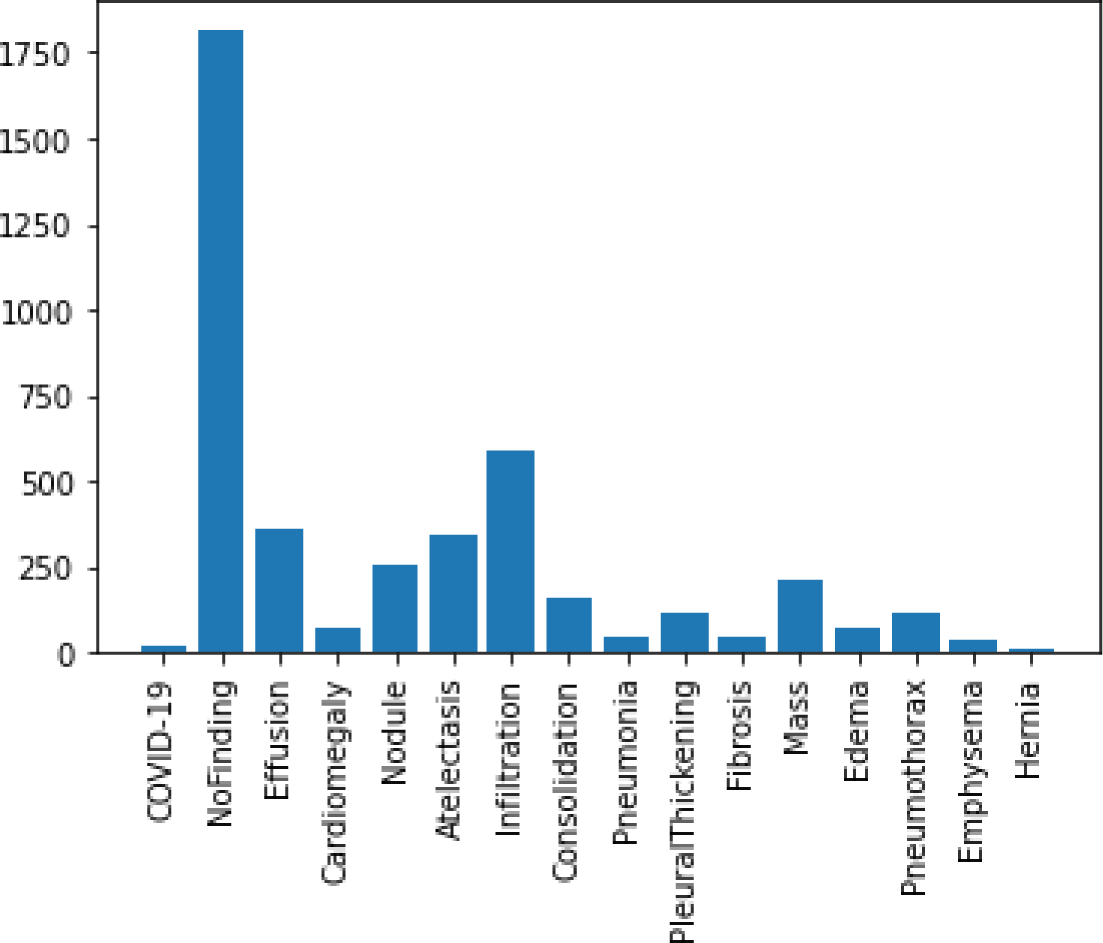
Distribution of testing samples among classes of disease as drawn from the Covid-19 Chest X-ray and NIH Chest X-ray datasets

Considering the level of noise, distortion or anomalies that might be associated with some of the images accessed from the public databases, this study attempted to pre-process all the samples. This was achieved by applying some standard image preprocessing techniques to our images. The next section details this approach.

### 2.2 Image Preprocessing

To enhance the performance of deep learning models, several studies [12, 13, 14, and 15] have encouraged the application of inputs/samples to appropriate preprocessing techniques/algorithms. Basically, image processing, which is the use of algorithms to perform image processing on digital images, is categorized into two: namely, the analogue image processing and digital image processing. The preprocessing techniques are aimed at improving the features in the image through image enhancement and the suppression of unwanted distortions thereby yielding an improved image/input for the deep learning model. In this study, we applied our samples to the following preprocessing techniques after reading or loading images into the buffer/memory:

- Image resizing: Due to the heterogeneity of the databases and as a result of variation in the sizes of images, we resized the images into 220×220 sizes. Such resizing operation allows for decreasing the total number of pixels from 888×882 and 1024×1024 for COVID-19 X-ray and NIH Chest X-Ray datasets respectively to 220×220 for both.
- Removal of noise (denoise): Image denoising can present a challenging procedure arising from the operation of estimation of the original image through the elimination of noise from a noisy image. For instance, one might be interested in removing any of the following noises from an image: Poisson noise, salt and pepper noise, Gaussian noise, and speckle noise. In this study, we attempted to eliminate/remove noise from our image samples using the Gaussian Blur technique since study [16] showed that the technique is relevant in images with high noise. We used a Gaussian filter by applying our images to the function cv2.GaussianBlur using kernel size of 5×5 and zero (0) for both the standard deviation for both the x and y directions.
- Morphology (smoothing edges): As a preprocessing operation before applying segmentation to our images, we applied the morphology operation to our samples. This enables us to extract image components that are useful in the representation and description of region shape. This operation (morphological smoothing) was aimed at removing bright and dark artifacts of noise, and was achieved through an opening followed by a closing operation. The output of this phase yielded images whose edges are smoothened for easy detection.
- Segmentation: It is well known that image segmentation allows for the partitioning of an image into multiple image objects or segments appearing as different categories of pixels such that a similar category constitutes a segment. This study approached this technique with the aim of enhancing the process of detecting image objects which support feature extraction thereby obtaining meaningful results. We achieved this through the application of thresholding method leaving out other methods such as edge detection based techniques, region based techniques, clustering based techniques, watershed based techniques, partial differential equation based and artificial neural network based techniques. Using the thresholding method, we THRESH_BINARY_INV thresholding style of opencv, and a maxVal of 255 which represents the value to be given if pixel value is more than (sometimes less than) the threshold value. The computation of THRESH_BINARY_INV is as shown in equation 1.

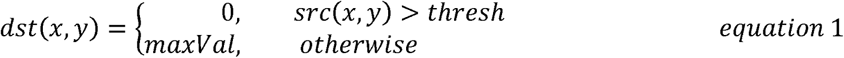

The second parameter to the maxVal is the retVal as used in our thresholding technique. We used Otsu’s method which is widely reported to yield interesting results and is also suitable when there is distinguishable foreground and background [17]. The use of this method is inferred from the value we set for the retVal which is the THRESH_OTSU. This allows for automating the process of calculating the threshold value from image histogram. Thus far, we have filtered our image samples with a 5×5 Gaussian kernel to remove the noise, and then applied Otsu thresholding. Furthermore, we applied the dilate operation on the image to enlarge the foreground and thereby find the sure background area. Also, to find the sure background area in the image, we applied the distanceTransform operation to achieve a representation of a binary image so that the value of each pixelwas replaced by its distance to the nearest background pixel. Hence the threshold segmentation was applied to divide the image into regions of object and background. Our thresholding segmentation was completed through the application of global thresholding which uses any appropriate threshold value of T= kept constant for the whole image so that the output image is obtained from original image as seen in equation 2 [18].

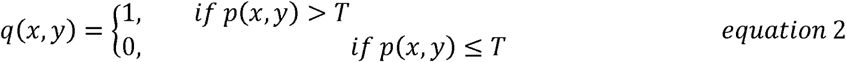

The resulting images from all the preprocessing techniques above are then passed as input into the CNN model described in subsection 3.3.

### 2.3 Detection and Classification Model for Covid-19

The proposed CNN model is a fraction of a complete framework in Figure 8 which represents the pipeline flow of techniques used in this study. The architectural pipeline shown in the figure below first reads in the sample images from the buffer where the combined datasets from the two databases are stored. Thereafter, the preprocessing techniques described in Subsection 3.2 are applied sequentially on them. Furthermore, the resized and improved image samples are split into training and validation sets based on the illustration shown in Subsection 2.1. Thereafter, the CNN model is applied to the input samples for training and validation. The trained model is then exposed to the testing set of images for prediction and then the result of the classification is output for performance evaluation.

**Figure 8:**
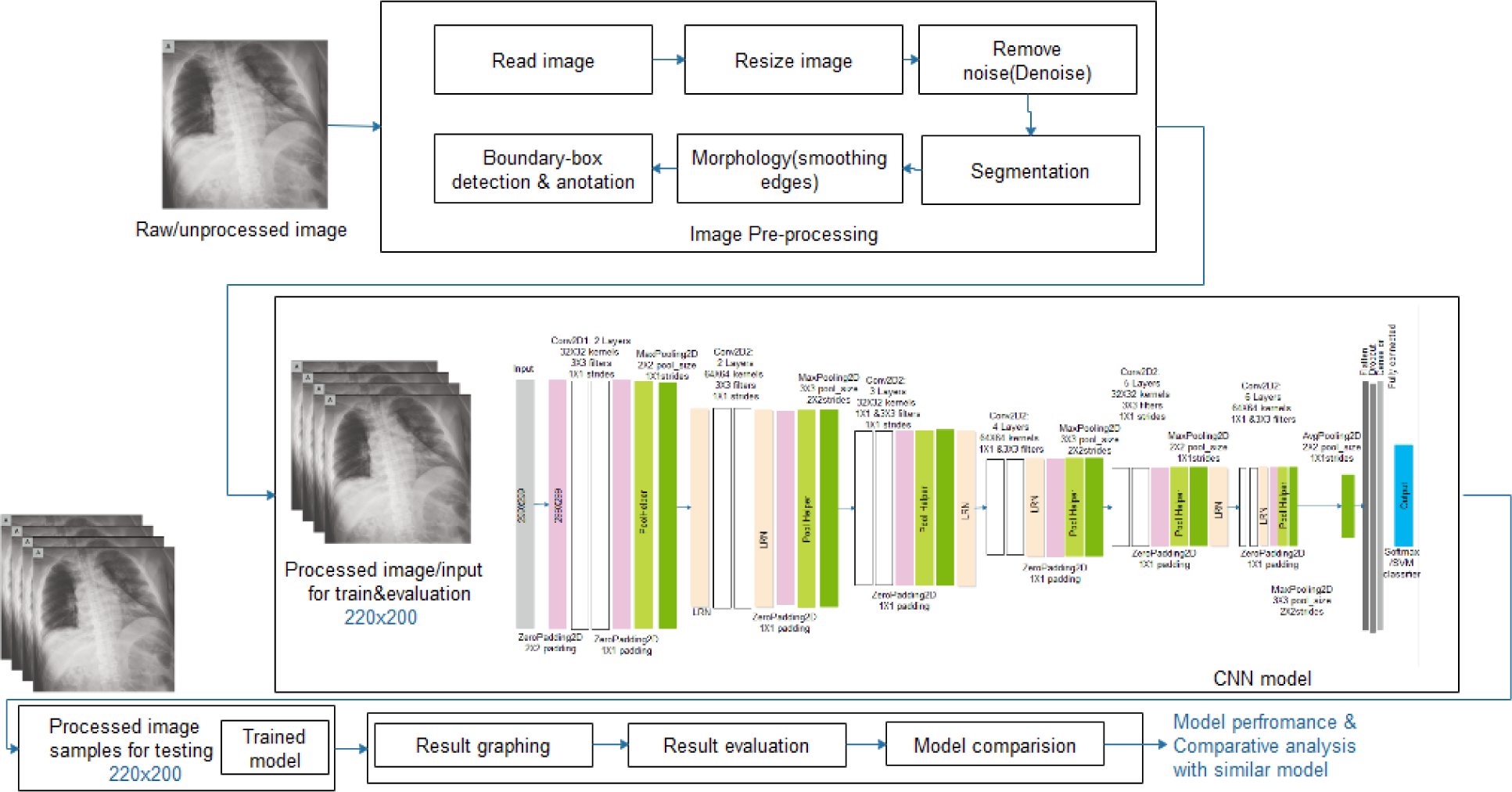
The architectural pipeline of the proposed image/input pre-processing, feature detection, sample classification and post-classification processing

CNN is the most widely used deep learning model for image recognition. Medical image recognition tasks have largely benefited from the fielding of computing. CNN consists of a convolution layer that extracts the features of the image and a fully connected layer that determines which class the input image belongs to – classification of the input image. In Figure 9, we present the architecture of the proposed CNN model designed and applied to our datasets in this study. The architecture of the model follows the form of Conv-Conv-Pool-Drop-Conv-Conv-BatchNorm-Pool-Drop-Dense(relu)–BatchNorm-Drop, with a number of filter modeled as 32(3, relu)–32(3, relu)-2(2)-64(3, relu)–64(3, relu) and so on. For the classification purpose, we applied the SoftMax function to the output of the feature detection phase of the model. This allows for a multiclass classification as against the binary classification in our case.

**Figure 9:**
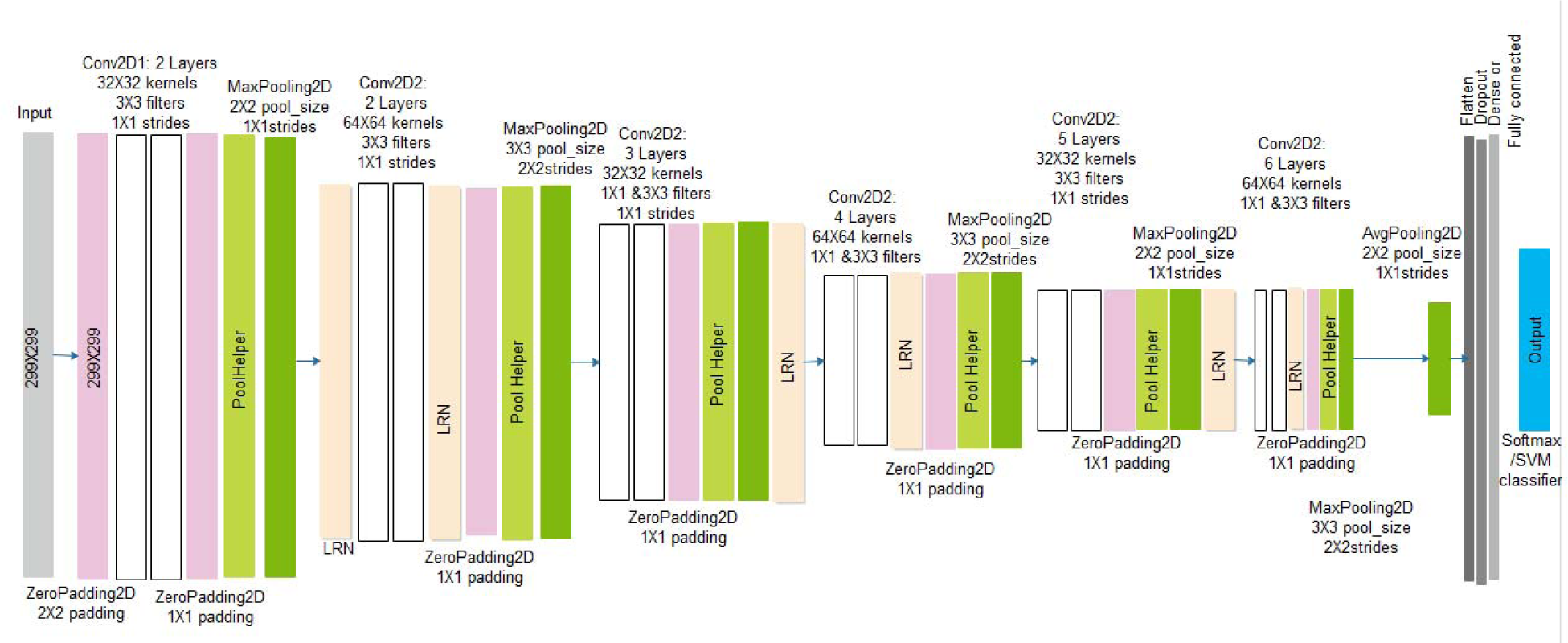
The architecture of the proposed convolutional neural network (CNN) for feature detection and classification Covid-19 disease from chest images

The proposed CNN model benefits from some deep learning regularization techniques which have demonstrated capacity to combat overfitting issue. Overfitting is the situation when a model learns the training data excellently but falls short of generalizing well when some other data is exposed to it. Regularization techniques such as L2 and L1, dropout, data augmentation, and early stopping have been widely reported to enhance the performance of deep learning models [19, 20]. This study therefore experimented with some of the techniques to ensure an optimal performance of the proposed deep learning (CNN) model. Hence, we do not just hope to improve performance but enable our model to generalize well. A model failing to generalize well will show validation error increases while the training error steadily decreases. In this study, we applied our work to the most common regularization technique L2 which is also referred to as “*weight decay*”. We aimed at applying this weight regularization technique to reduce overfitting. L2 values ranges between 0 and 0.1 with examples as 0.1, 0.001, 0.0001, and are in logarithmic scale. We therefore hoped to reduce our model’s training error [21, 22] by applying this technique. For instance, the Inception V3 model experimented with a value of 0.00004 and we discovered that it was suboptimal and instead experimented with 0.00005 [23]. In addition to the use of L2, we also demonstrated the use of early stopping to stop our model from continuing training when it had attained its optimal performance. This regularization concept is another widely used technique in deep learning to stop training when generalization error increases. In addition to the use of data augmentation technique, the proposed CNN model was also allowed to use dropout layer at the rate of 0.5.

## 3 Experimentation

In this section, the COVID-19 chest X-ray and NIH chest X-ray datasets described in subsection 2.1 are trained based on our CNN model and the performances of multiclass classification are evaluated. The environment for the experimentation and the outcome of the preprocessing techniques are also described in this section.

### 3.1 Implementation Environment

All our experiments were carried out on Google’s Colab environment with the following configurations as the need arose: 2-core Intel(R) Xeon(R) CPU @ 2.30GHz, 13GB memory and 33GB hard drive; and GPU Tesla P100-PCIE-16GB.

### 3.2 Image Pre-processing

The pre-processing techniques applied to our input images/samples were extensively discussed in subsection 2.2. Therefore we aim to present the outcome of the application of those techniques on our datasets. The first operation applied was the resizing of images from the high resolution of 888×882 and 1024×1024 to a collective size of 220×220. This was necessary to allow the datasets sourced from different platforms to feed into our model effectively as fixed size. In Figures 10 and 11, we show the original image samples from Covid-19 and the NIH chest X-ray datasets respectively, and the outcome of the resizing of the operation is shown in Figure 12.

**Figure 10:**
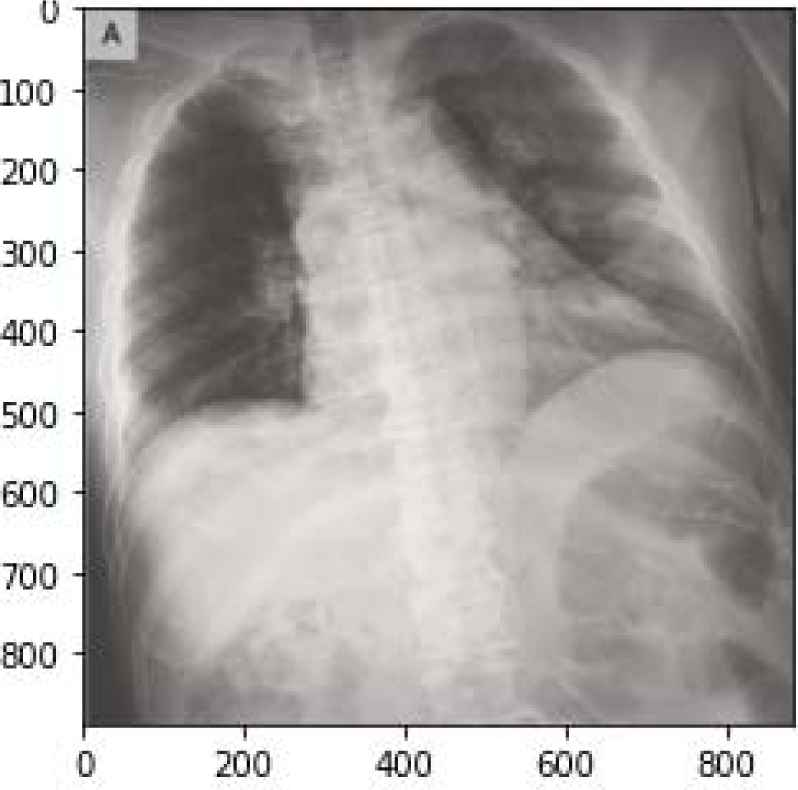
A sample of original/raw image of size 888×882 from the Covid-19 Chest X-ray dataset

**Figure 11:**
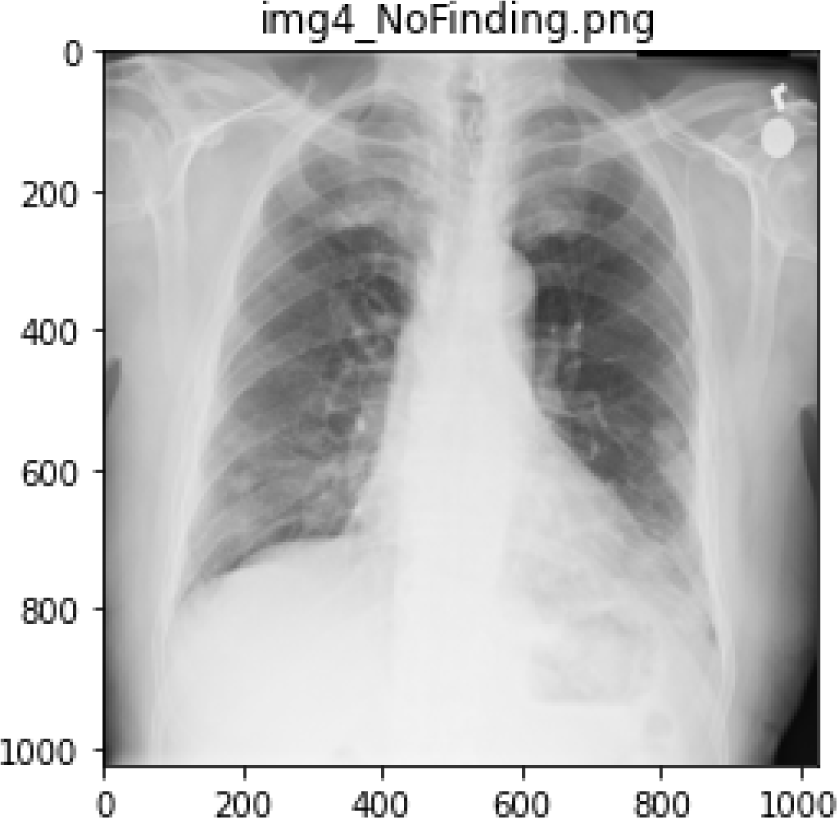
A sample of raw image labeled ‘No Finding’ of size 1024×1024 from the NIH Chest X-ray dataset

**Figure 12:**
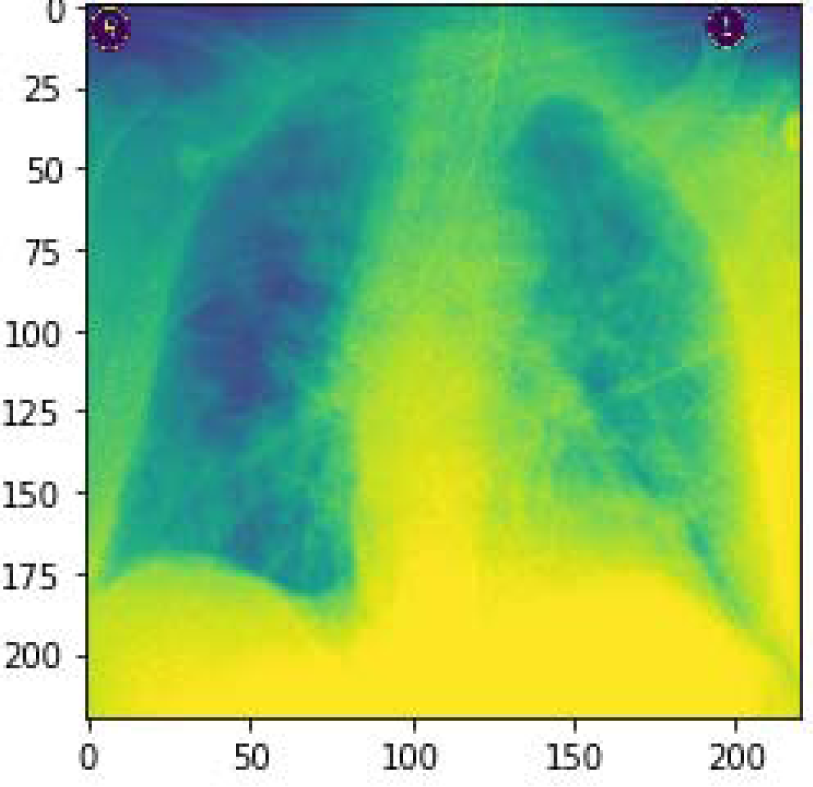
A resized sample from 888×882 to 220×200 in the Covid-19 Chest X-ray dataset

One major pre-processing operation carried out on our input sets is the removal of noise as described in subsection 2.2. The approach taken in this study is to blur the image samples as a measure to clean and denoise it. Hence, in Figure 13, a pair of samples resulting from an un-denoised and then denoised image are captured and shown. Furthermore, to demonstrate the segmentation operation carried out on the image samples by this study, we have also presented output from such operation. In Figures 14 (a-b), a pair of samples of images whose segments and background are extracted are presented. The pair of images in Figure 14a shows the original image and the outcome of the segmented image, while that of Figure 14b shows the original image and its extracted background. These operations allow for easier understanding of what is in the image and enable easier analysis of each part. In addition, the segmentation operation on our medical X-ray images revealed objects which were unknown, but were segmented within the image typically for further investigation.

**Figure 13:**
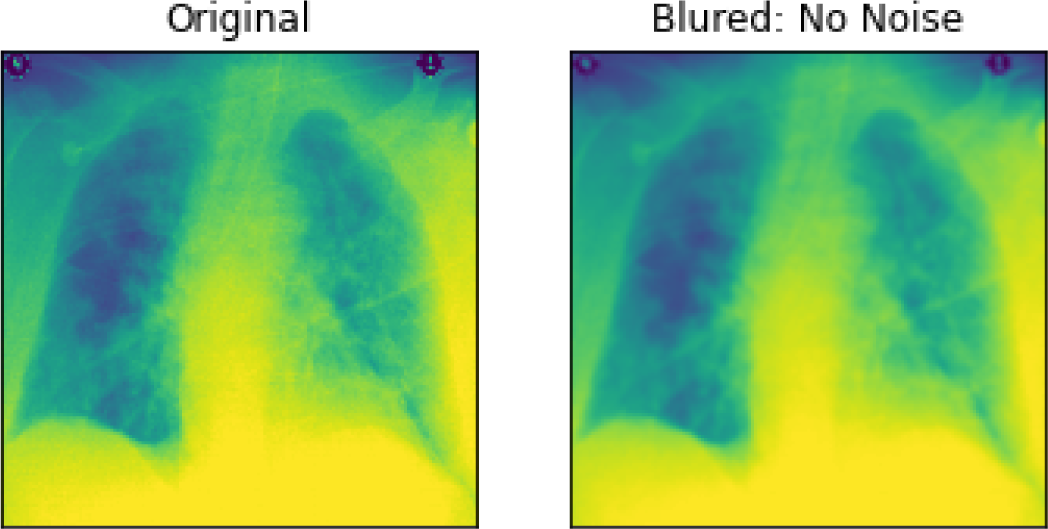
An illustration of image pre-processing task carried out on samples through the denoising and blurring effect

**Figure 14a:**
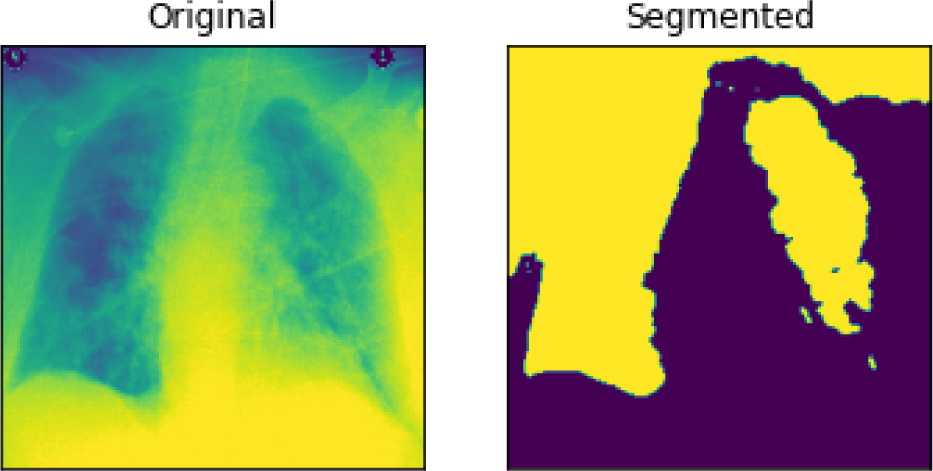
An illustration of image pre-processing task aimed at segmenting samples

**Figure 14b:**
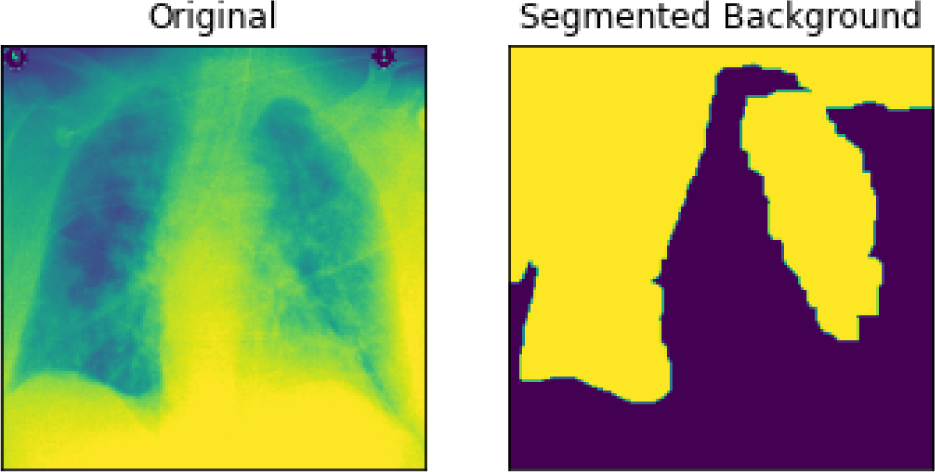
An illustration of image pre-processing task aimed at extracting segmented background from samples

The use of bounding boxes is one of the most interesting operations that provide support for image annotation in deep learning models. This proves useful in object classification in images and even for further localization tasks. Whereas image is aimed at assigning a class label to an image, object localization allows for creating bounding boxes around recognizable objects in the image. The target of the model to classify and also obtain positions of objects in image is referred to as object detection or object recognition. Drawing bounding boxes can be achieved using deep learning models or other algorithms. For instance, to describe the location of some targeted diseases in our input images, we draw a bounding box as a rectangular box that can be determined by the x and y axis coordinates in the upper-left corner and the x and y axis coordinates in the lower-right corner of the rectangle. This operation allows for easily annotating our samples for convenient recognition by CNN model.

Figures 15 (a-b) and 16 (a-b) show the bounding boxes (using black color) locating the position of the labeled disease on the image and their corresponding contours. In addition to drawing bounding boxes around input, we detected contours both in the bounding box and those outside it. In each case of images in Figures 15 and 16, the upper image represents the bounding box localizing presence of the disease while the lower figure represents the contours in each image.

**Figure 15:**
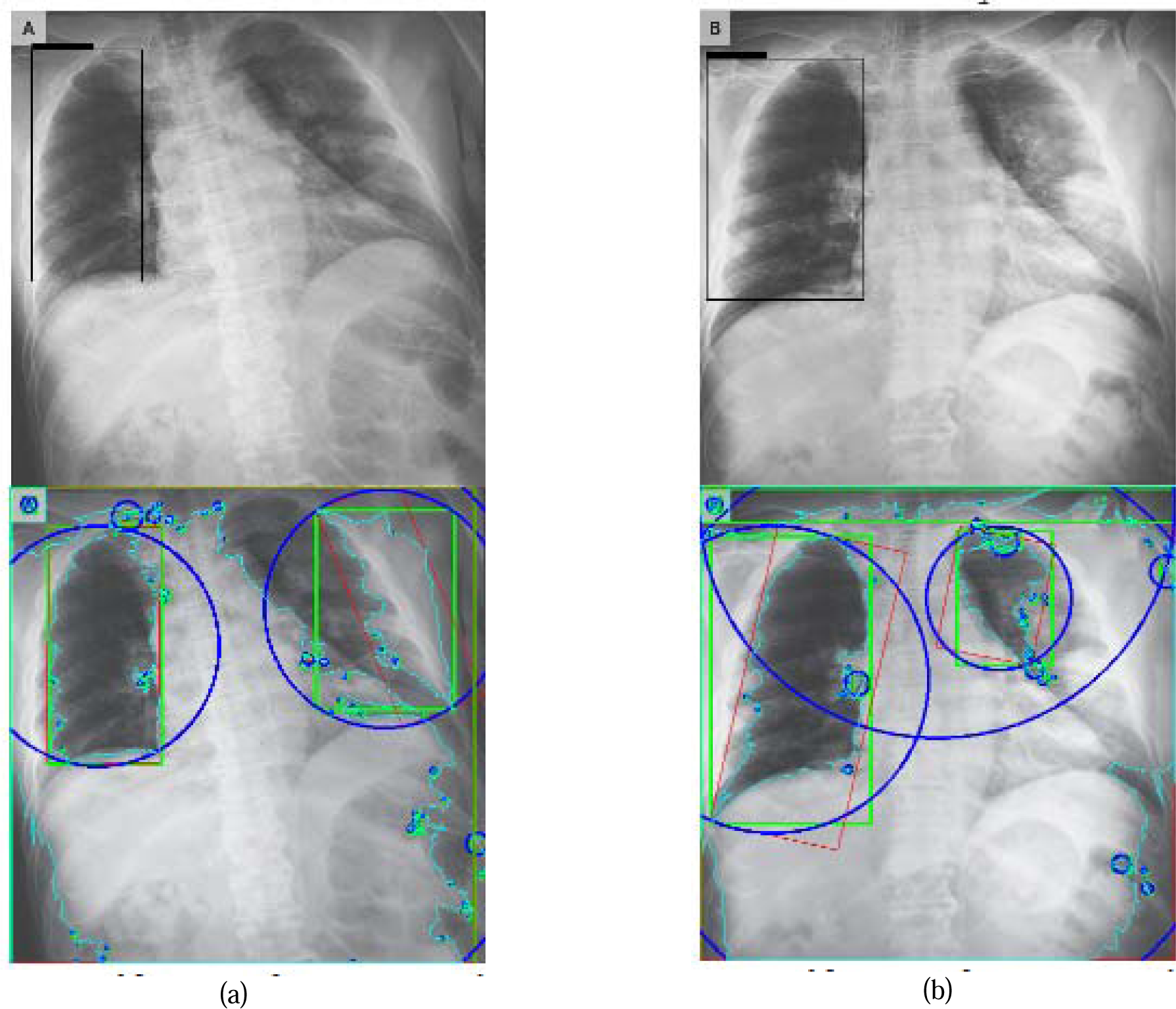
Samples of images and their respective findings with expert annotation as extracted from the NIH Chest X-ray dataset. The bounding boxes show the suspected regions of COVID-19 in PA view. Below is a capture of the contours in the image samples.

**Figure 16:**
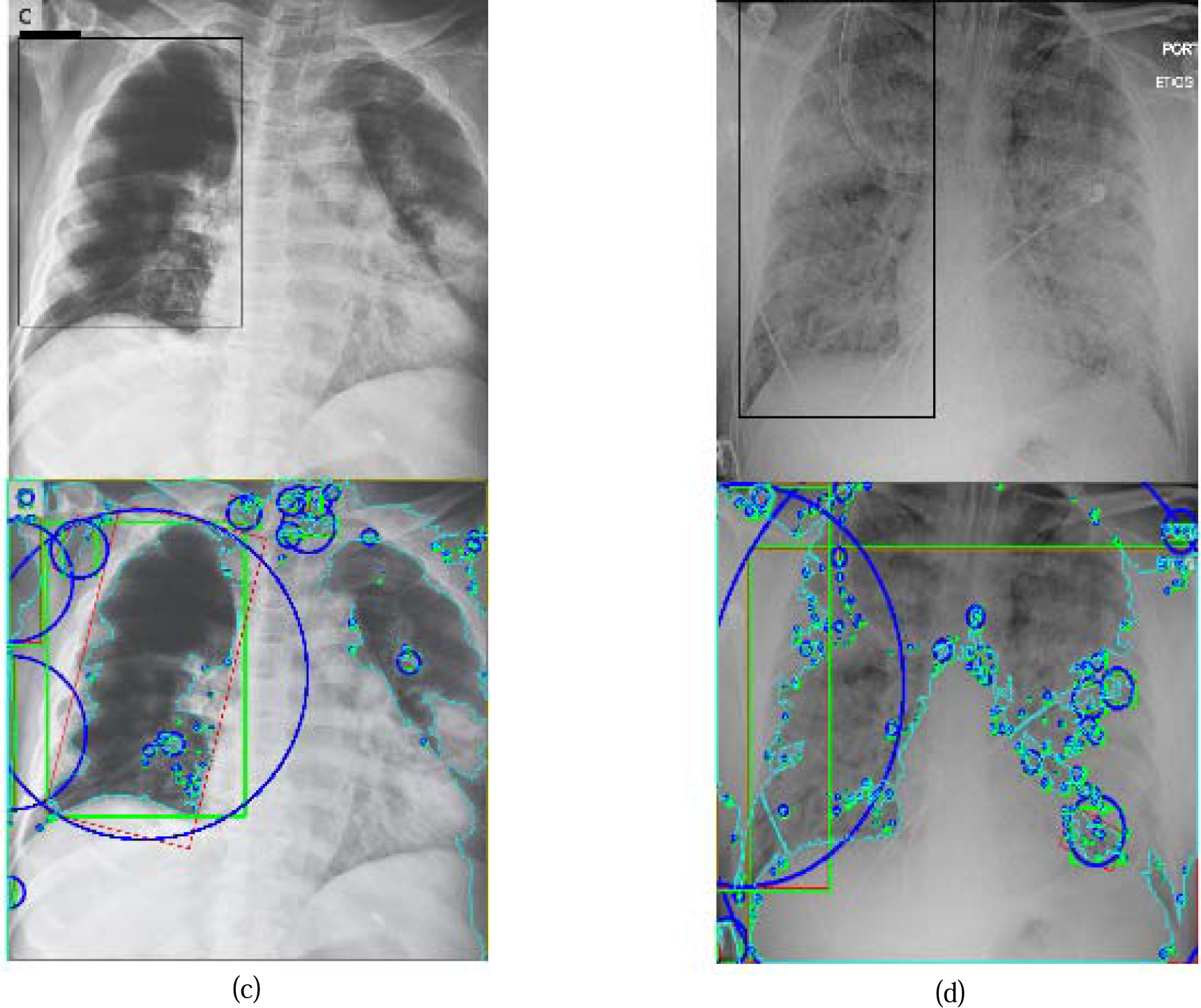
Samples of images and their respective findings with expert annotation as extracted from the NIH Chest X-ray dataset. The bounding boxes show the suspected regions of COVID-19 and ARDS in PA views. Below is a capture of the contours in the image samples.

Contours allow for the identification of the shapes of objects within an image, and are recognized through lines of curves joining all the continuous points with similar color or intensity. This technique provides support for object detection and recognition. In this study, to extract the contours as shown in the images below those with the bounding boxes, we carry out the following: we first threshold each image and then find all the contours in each image; with each contour, we draw a bounding rectangle in green color; then get a minimum area rectangle and convert all coordinates floating point values to integer, and draw a red ‘nghien’ rectangle; furthermore, we get the minimum enclosing circle and convert all values to integer to draw the circle in blue; then finally draw all contours on each image.

### 3.3 Experimental Results

The proposed CNN model receives grayscale images as its input and experiments are performed with multiclass classifications. Table 1 shows the detection classes for each classification and their distribution in both datasets. Meanwhile, for each experiment carried out, we train the model for 50 epochs and 1310 steps.

**Table 1:** Classes for multiclass classifications for the COVID-19 and National Institutes of Health (NIH) Chest X-ray datasets

| Class | COVID-19 (as chest X-ray or CT images) Multiclass | NIH Multiclass |
| --- | --- | --- |
| 1 | Covid-19 | Covid-19 |
| 2 | MERS | Atelectasis |
| 3 | SARS | Hernia |
| 4 | ARDS | Mass |
| 5 | No-Finding | No-Finding |
| 6 | Streptococcus | Consolidation |
| 7 | E.coli | Infiltration |
| 8 | Pneumocystis | Pneumothorax |
| 9 | Chlamydophila | Edema |
| 10 | Legionella | Emphysema |
| 11 | Klebsiella | Fibrosis |
| 12 |  | Effusion |
| 13 |  | Pneumonia |
| 14 |  | Pleural thickening |
| 15 |  | Cardiomegaly |
| 16 |  | Nodule |

We experimented with the proposed CNN model on the datasets using some variation of hyperparameters. For instance, we investigated the performance of the model when SGD and Adam optimizers were applied to the model and plotted the output of the model. Furthermore, we experimented on our proposed model the effect of two different values for the L2 (weight decay) regularization technique. Meanwhile, the proposed CNN model was also applied to classical data augmentation technique to determine possible improvement in its performance.

The first set of experiments used the Adam optimization algorithm and weight decay (L2) value of 0.0002. Figure 17 captures the performance of the model in terms of loss function while Figure 18 shows the trajectory of the accuracy for both training and validation cases. We plotted the confusion matrix of the experimentation after carrying out a prediction using the test datasets and this is shown in Figure 19. Note that the configuration of the Adam optimizer is as follows: learning rate=0.001, beta1=0.9, beta2=0.999 and epsilon=1e-8.

**Figure 17:**
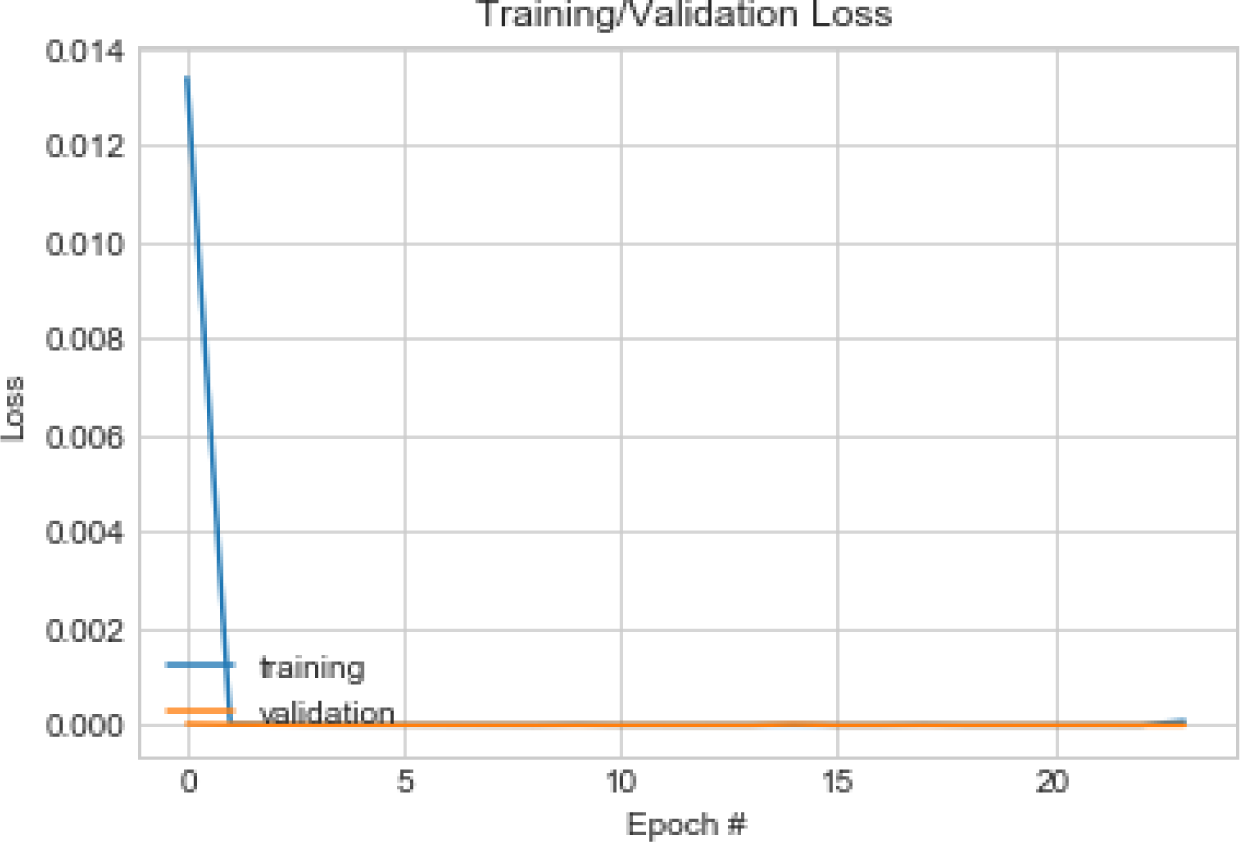
Pattern of change in loss function of training and validation on the combined dataset using Adam optimizer

**Figure 18:**
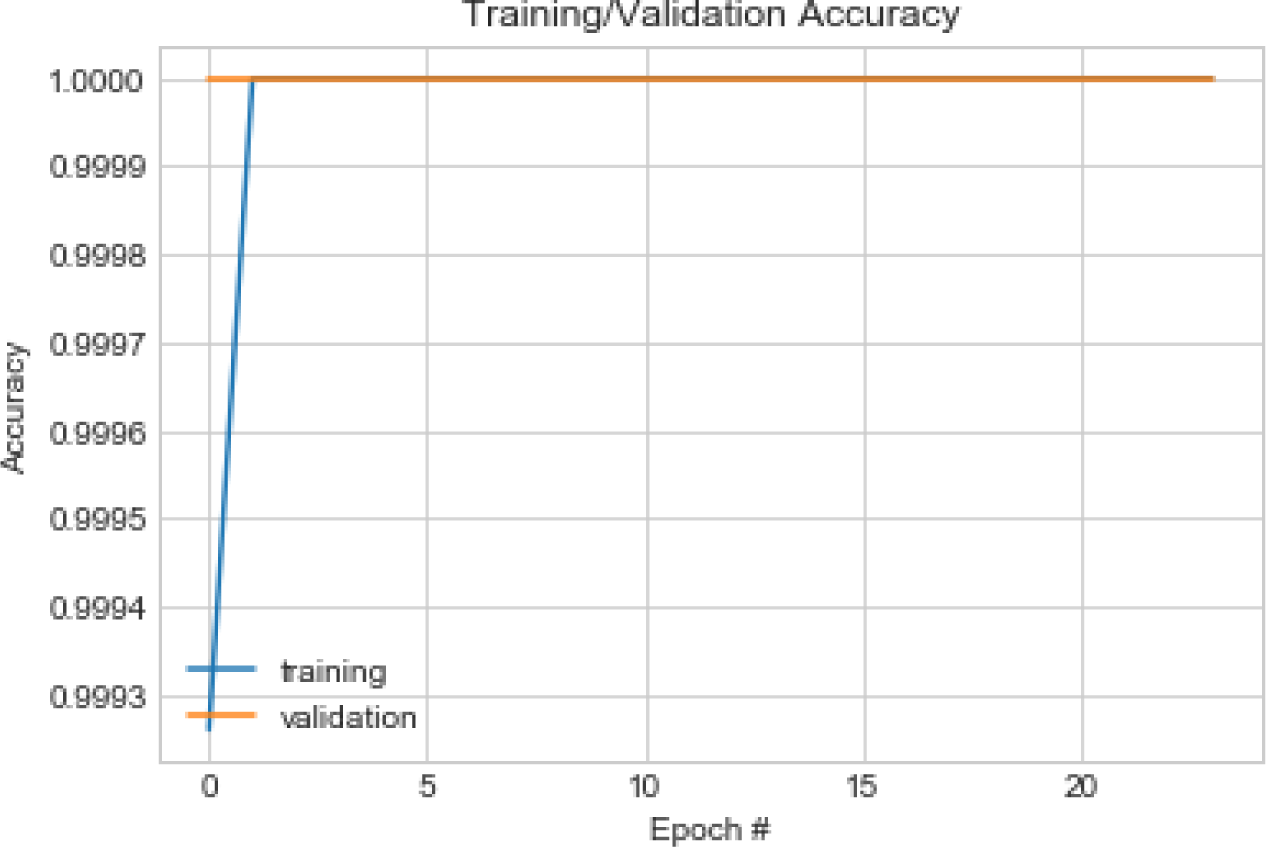
Pattern of change in accuracy of training and validation on the combined dataset using Adam optimizer

**Figure 19:** Confusion matrix showing the multi-class classification which includes COVID-19 against other lung diseases from the testing sets using Adam optimizer

Similarly, in the second experiment performed, we experimented using the SGD optimizer with the following configuration: learning rate=0.01, decay=1e-6, momentum=0.9 and nesterov=True. The value of 0.0005 was used for the L2 regularization technique. The performance of the model was also examined and we found that although the accuracy remained close to that of Adam optimizer, there was, however, a difference in the loss values trajectory. Figures 20 and 21 capture the performance of the model on the training and validation datasets in the cases of loss function and accuracy. We also exposed the trained model to the test dataset under the same configuration and plotted the confusion matrix which is shown in Figure 22.

**Figure 20:**
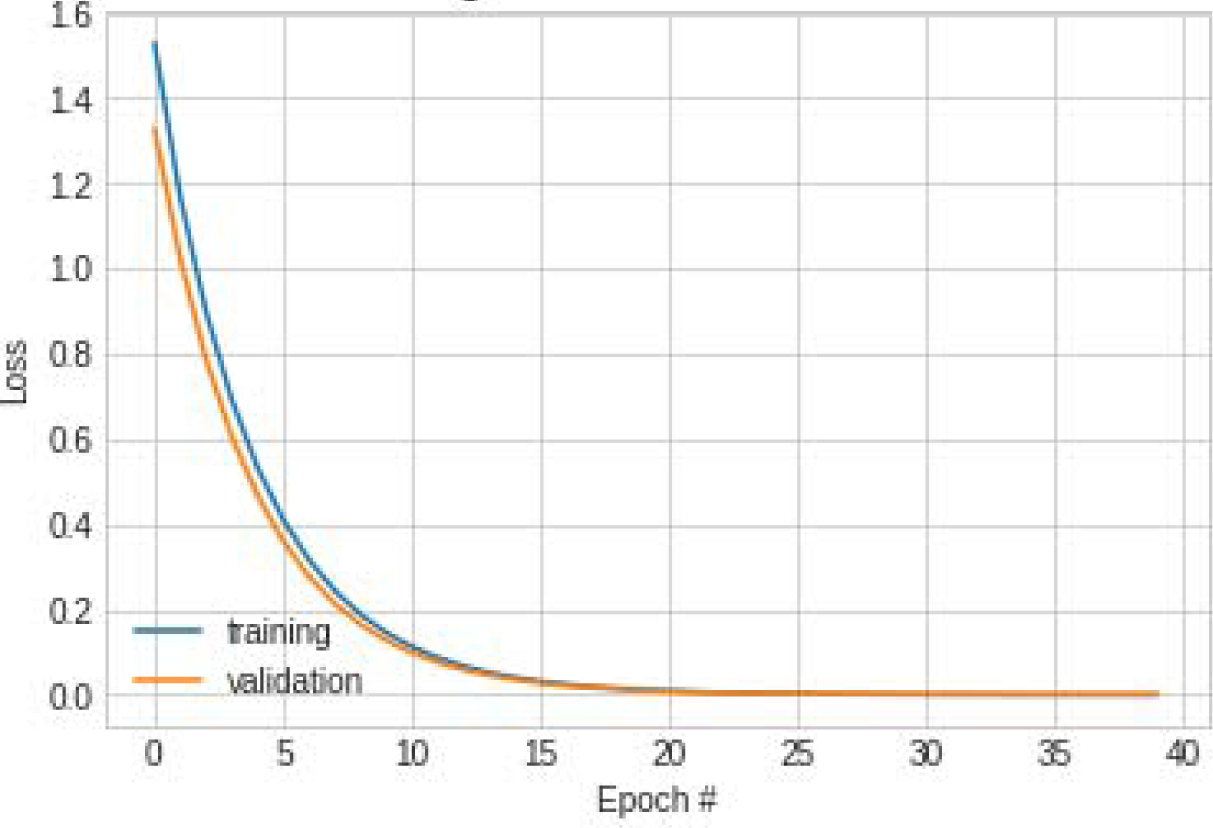
Pattern of change in loss function of training and validation on the combined dataset using SGD optimizer

**Figure 21:**
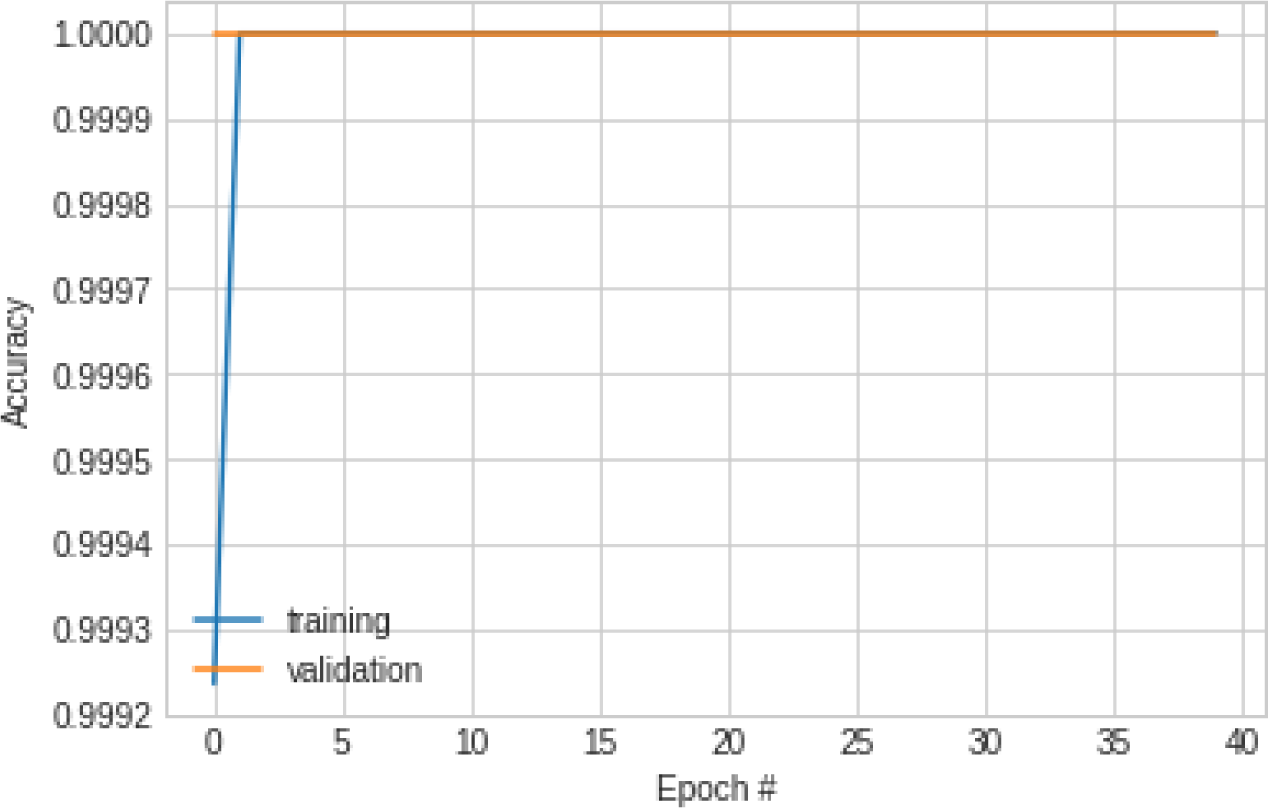
Pattern of change in accuracy of training and validation on the combined dataset using SGD optimizer

**Figure 22:** Confusion matrix showing the multi-class classification which includes COVID-19 against other lung diseases from the testing sets using SGD optimizer

The outcome of applying the confusion matrix operation on the output of the prediction yielded the values below across the classes (18) found in the test datasets. In Figure 22 we present a graphical representation for it.

In subsection 2.3, we noted that one of the regularization techniques applied to our proposed CNN model is the Early Stopping technique. We found this application useful as it helped to stop our model once it recognized that optimal performance was attained. We show evidence of this in Figure 23 where the training was terminated at 24^th^ epoch during the first experiment where data augmentation was not applied.

**Figure 23:**
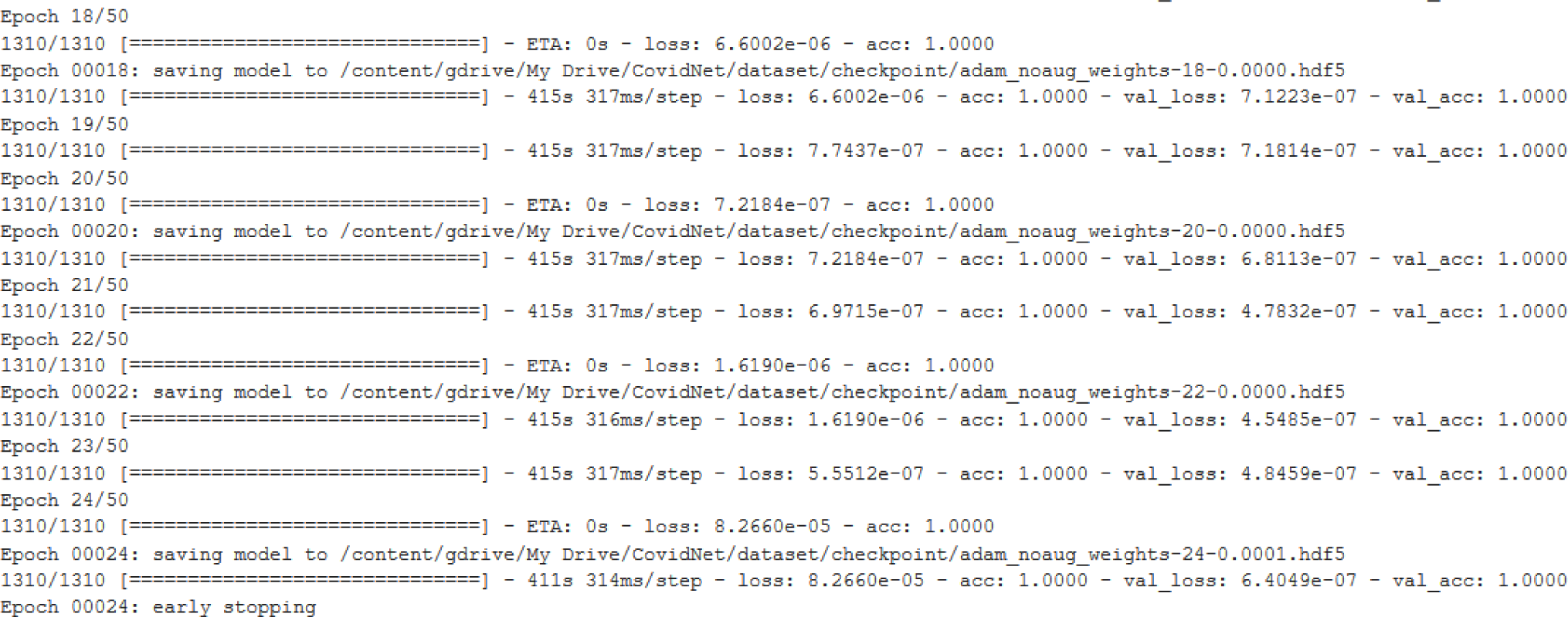
The outcome of the application of one of the regularization techniques Early Stopping as discussed in Subsection 2.3

Now that the experimentation on the proposed CNN model has been presented, we shall proceed to the next section to discuss the performance of the model compared with some state-of-the-art models diagnosing Covid-19 disease.

## 4 Results and Discussion

The experimental results of multiclass classification for four (4) experimental scenarios show that most scenarios have more than 99% accuracy. To evaluate the performance of the proposed model, we calculate accuracy, sensitivity, specificity, precision, recall, F1-score, Cohen’s Kappa, ROC AUC, and confusion matrix. In the following paragraphs, we briefly outline the metrics and their relevance to our classification of novel COVID-19 disease.

The computational metric precision checks what proportion or quantity of positive identifications achieved by a model were actually correct and is given by equation 3.

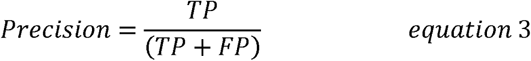

On the other hand, recall checks the number of actual positive cases in our datasets which the proposed CNN model was able to correctly identify. This is given by equation 4.

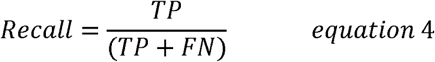

Evaluating the effectiveness of our CNN model requires that we examine its performance in terms of precision and recall, hence the need for the computation of these metrics. Furthermore, we examined another metric known as F1 Score. This metric expresses the balance between the precision and the recall described above and helps us decide whether the performance of our model is based on precision and recall. We give the equation for F1 score in equation 5.

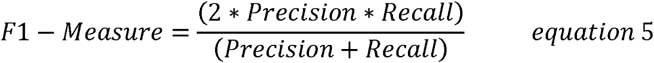

In this study, we chose an under-utilized, though effective at multiclass classification, metric known as Cohen’s Kappa. This metric is robust in handling imbalanced class problems as may be seen in our datasets. In a multiclass classification problem, this metric provides a wider view of the performance of a classification model compared to accuracy (in equation 6) or precision/recall. The metric is represented in equation 7.

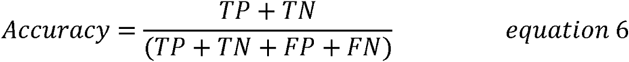

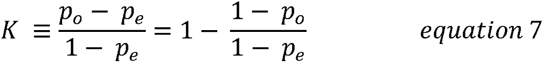

The receiver operating characteristic (ROC) curve expresses the performance of our classification (CNN) model using a graphical approach and does this at all classification thresholds. It is able to achieve this by graphing the True Positive Rate (TPR) and False Positive Rate (FPR). The metric gives a summary of the performance of a classifier over all possible thresholds. Similar to the ROC is the area under the ROC curve (AUC) which examines the entire two-dimensional area underneath the entire ROC curve which covers from (0,0) to (1,1). This metric is effective at checking the proper/wellness and quality of our model’s prediction performance.

Finally for the description of our metrics, we have the confusion matrix. Whereas accuracy of a model may seem appealing in some sense, it is, however, limited by its inability to give detail of the performance of the classification model. On the other hand, confusion matrix presents this detail by presenting the prediction result in an unambiguous manner.

In Table 2, we list the performance of our model for the experiments carried out in comparison with similar models adapted to the purpose of classification Covid-19 disease.

**Table 2:** Summary of result obtained by the proposed model

| Authors | Contribution/<br>Approach | Specificity | Sensitivity | Precision/Recall | F-Score | AUC | Accuracy |
| --- | --- | --- | --- | --- | --- | --- | --- |
| Proposed approach<br>[Experiment 1:<br>Using Adam] | CNN for<br>classification<br>of Covid-19 | 1.00 | - | 0.85/0.85 | 0.90 | 0.50 | 1.00 |
| Proposed approach<br>[Experiment 2:<br>Using SGD] | CNN for<br>classification<br>of Covid-19 | 1.00 | - | 0.85/0.85 | 0.90 | 0.50 | 1.00 |
| Tartaglione et al [8] | COVID-Net | 0.80 | 0.90 | - | 0.90 | 0.85 | 0.85 |
| Ko et al [10] | FCONet | 0.100 | 0.9958 | - | - | - | 0.99 |

The result obtained in Table 2 shows that our system achieves 1.00, 0.85, 0.85, 0.90, 0.50, and 1.00 for specificity, recall, and precision, F-score, AUC and accuracy respectively in phase one of the first experiment. On the other hand, in the second experimentation we carried out, our model yielded the following achieves: 1.00, 0.85, 0.85, 0.90, 0.50, and 1.00 for specificity, recall, and precision, F-score, AUC and accuracy respectively in the phase one of the second experiment. The proposed model attains an 85% value for both precision and recall which made it useful for the proposed task, hence avoiding unnecessary false alarms. **F1 measure** is relevant if we are looking to select a model based on a balance between precision and recall, and is the harmonic mean of precision and recall and gives a better measure of the incorrectly classified cases than the accuracy. As a result, the value of 0.9 for our F1-score shows the performance of our model even when there are imbalanced classes as is the case in most real-life classification problems.

Confusion matrix is well known as an unambiguous way to present prediction results of a classifier and works for both binary and multiclass classification. When binary classification is desired, confusion matrix is usually presented in a 2×2 table showing true positive, true negative, false positive, and false negative. However, in this study which aims at multiclass classifications, the table has a size equal to the number of classes (24) squared. The confusion matrix computed by our proposed model using both Adam SGD (optimizers) in making predictions of different classes in the combined dataset is shown in Figures 19 and 22. Generally, it is known that the higher the number on the diagonal of the confusion matrix, the better the accuracy of the model. Evidently, our proposed model achieved a good performance with respect to this. One of the most useful metrics is the classification_report which combines several measures and prints a table with the results.

Further to the presentation and comparison of the performance of our model using metrics like specificity, sensitivity, recall, precision, F-score, AUC and others, we use Table 3 to show the performance of our model with others in terms of accuracy.

**Table 3:** Comparing the contributions and performances of the proposed study with similar approaches

| Authors | Contribution/Approach | Dataset | Accuracy |
| --- | --- | --- | --- |
| **Proposed approach** | **Deep learning architecture for effective classification of Covid-19** | **Covid-19 and NIH Chest X-ray datasets** | **100.00%** |
| Apostolopoulos et al [6] | VGG19, Mobile Net, Inception, Xception, and InceptionResNet v2 |  | 93.48% |
| Ozturk et al. [4] | DarkCovidNet | Raw chest X-ray images | 87.02%, |
| Khan et al. [7] | CoroNet |  | 89.60% |
| Wang et al. [5] | COVID-Net |  | 92.40 % |
| Tartaglione et al [8] | COVID-Net and ResNet-18 | COVID-ChestXRay dataset | 85.00% and 100.00% respectively |
| Yoo et al [9] | Deep learning-based Decision-tree classifier | COVID-ChestXRay dataset | 95% |
| Ko et al [10] | fast-track COVID-19 classification network (FCONet) | 3993 Chest CT images privately collected | 99.87% |
| Panwar et al [11] | nCOVnet | Chest X-Ray Images | 97.00% |

Considering the performance of our model from Table 3 compared with other similar studies, we conclude that this study outperforms state-of-the-art deep learning models aimed at detecting and classifying the novel Coronavirus (Covid-19). This is readily seen from the performances of each study. We note that only Ko et al [10] and Panwar et al [11] who used fast-track COVID-19 classification network (FCONet) and nCOVnet respectively have their models’ performances competing with our model. This study has, therefore, successfully advanced research in the areas of detection and classification of COVId-19 using deep learning models.

## 5 Conclusion and Future Work

In this paper a deep learning model based on CNN was designed and implemented for the purpose of detecting and classifying the presence of COVID-19 in chest X-rays and CT images. We applied the proposed CNN model to two publicly available datasets, namely COVID-19 chest X-ray and the NIH chest X-ray databases. In addition, we enhanced the performance of our model by applying it to some regularization techniques. Furthermore, we investigated the performance of the proposed model by juxtaposing the use of optimizer between the popular Adam and SGD. The result obtained revealed that our model achieved 100% accuracy in classifying the novel coronavirus (COVID-19). With the exponential increase in COVID-19 reported cases and treatments around the globe, the volume of COVID-19 datasets is being created and archived daily. Therefore, future studies could focus on advancing the architecture of the proposed deep learning model presented in this paper and most importantly investing the robustness of this model on some large scale datasets. Furthermore, it will also be interesting to see the deployment of our trained CNN based deep learning model to both web and Android applications for clinical utilization.

## Data Availability

Data are publicly available on chest X-ray databases

